# Rapid, large-scale wastewater surveillance and automated reporting system enabled early detection of nearly 85% of COVID-19 cases on a University campus

**DOI:** 10.1101/2021.06.18.21259162

**Authors:** Smruthi Karthikeyan, Andrew Nguyen, Daniel McDonald, Yijian Zong, Nancy Ronquillo, Junting Ren, Jingjing Zou, Sawyer Farmer, Greg Humphrey, Diana Henderson, Tara Javidi, Karen Messer, Cheryl Anderson, Robert Schooley, Natasha Martin, Rob Knight

**Affiliations:** Department of Pediatrics, School of Medicine, University of California, San Diego, La Jolla, California, USA; Herbert Wertheim School of Public Health and Human Longevity Science, University of California, San Diego, La Jolla, California, USA; Department of Urban Studies and Planning, University of California, San Diego, La Jolla, California, USA; Halicioglu Data Science Institute, University of California, San Diego, La Jolla, California, USA; Department of Electrical and Computer Engineering, University of California, San Diego, La Jolla, California, USA; Campus Planning, University of California, San Diego, La Jolla, California, USA; Division of Hypertension and Nephrology, School of Medicine, University of California, San Diego, La Jolla, California, USA; Division of Infectious Diseases and Global Public Health, School of Medicine, University of California, San Diego, La Jolla, California, USA; Department of Bioengineering, University of California, San Diego, La Jolla, CA, USA; Department of Computer Science & Engineering, University of California, San Diego, La Jolla, CA, USA; Center for Microbiome Innovation, University of California, San Diego, La Jolla, CA, USA

## Abstract

Wastewater based surveillance has gained prominence and come to the forefront as a leading indicator of forecasting COVID-19 infection dynamics owing to its cost-effectiveness and its ability to inform early public health interventions. A university campus could especially benefit from wastewater surveillance as they are characterized by largely asymptomatic populations and are potential hotspots for transmission that necessitate frequent diagnostic testing. In this study, we employed a large-scale GIS (Geographic information systems) enabled building-level wastewater monitoring system associated with the on-campus residences of 7614 individuals. Sixty-eight automated wastewater samplers were deployed to monitor 239 campus buildings with a focus on residential buildings. Time-weighted composite samples were collected on a daily basis and analyzed within the same day. Sample processing was streamlined significantly through automation, reducing the turnaround time by 20-fold and exceeding the scale of similar surveillance programs by 10 to 100-fold, thereby overcoming one of the biggest bottlenecks in wastewater surveillance. An automated wastewater notification system was developed to alert residents to a positive wastewater sample associated with their residence and to encourage uptake of campus-provided asymptomatic testing at no charge. This system, integrated with the rest of the “Return to Learn” program at UC San Diego-led to the early diagnosis of nearly 85% of all COVID-19 cases on campus. Covid-19 testing rates increased by 1.9-13X following wastewater notifications. Our study shows the potential for a robust, efficient wastewater surveillance system to greatly reduce infection risk as college campuses and other high-risk environments reopen.

**IMPORTANCE:** Wastewater based epidemiology can be particularly valuable at University campuses where high-resolution spatial sampling in a well-controlled context could not only provide insight into what affects campus community as well as how those inferences can be extended to a broader city/county context. In the present study, a large-scale wastewater surveillance was successfully implemented on a large university campus enabling early detection of 85% of COVID-19 cases thereby averting potential outbreaks. The highly automated sample processing to reporting system enabled dramatically reduced the turnaround time to 5h (sample to result time) for 96 samples. Furthermore, miniaturization of the sample processing pipeline brought down the processing cost significantly ($13/sample). Taken together, these results show that such a system could greatly ameliorate long-term surveillance on such communities as they look to reopen.

## INTRODUCTION

The recent SARS-CoV-2 pandemic has revealed the imminent need for rapid surveillance at the community level to track potential outbreak clusters ahead of clinical diagnosis, particularly considering the important role of asymptomatic infection in transmission. Wastewater based epidemiology has been extensively employed to monitor several enteric viruses such as polio across several global settings (1, 2). Wastewater monitoring has additionally been used as a surrogate to track the extent and spread of SARS-CoV-2 in communities - particularly when diagnostic testing is limited. Previous work has shown the ability of wastewater-based surveillance to foreshadow trends in diagnosis and hospitalization rates ahead of clinical testing, thereby serving as a barometer of the community infection dynamics (3–5). By providing an indicator of disease burden/prevalence in a given community, wastewater monitoring can act as an early warning system; potential clusters can be advised to undergo diagnostic testing increasing the opportunities for early public health interventions for individuals testing positive for SARS-CoV-2. Clinical diagnostics in tandem with wastewater-based surveillance systems can potentially provide a balanced community-wide perspective by increasing the chances of capturing both symptomatic and asymptomatic individuals. Wastewater-based surveillance can be particularly valuable at university campuses (which often self-maintain quasi-closed sewer systems) where, the detection of virus in wastewater in buildings where one or more infected individuals are living or working, can prompt testing of residents/employees in the affected buildings to diagnose and isolate individuals early on in their course of infection and avert outbreaks. With over 700,000 cases (https://www.nytimes.com/interactive/2021/us/college-covid-tracker.html) linked to university campuses in the United States alone, environmental or wastewater based surveillance can alleviate the burden of frequent diagnostic testing by focusing testing effort on potential hotspots (6). Tracking SARS-CoV-2 signatures in sewage at the building-level could enable targeted response strategies. Furthermore, high-resolution spatial sampling in a well-controlled context, coupled with detailed building use and occupancy data in a systematic manner integrated into a campus GIS network, can greatly augment data analytics (for instance, utilizing historical data to forecast high risk locations). In addition to the overarching social consequence for robust and rapid identification of SARS-CoV-2 outbreaks on college campuses, the university setting can also serve as an ideal test for collating and relating information about SARS-CoV-2 incidence and spread, which can facilitate similar studies across different scales and different types of communities. Over the last few months around 210 universities nationally and globally have implemented wastewater surveillance as a part of their SARS-CoV-2 monitoring programs (https://www.covid19wbec.org) (7–9). However, most of these campuses monitor either a few buildings or on an infrequent scale and none monitor on a daily basis.

The University of California San Diego (UCSD) implemented a multifaceted adaptive approach as a part of its Return to Learn (RTL) program, which aimed to improve campus safety through three interdependent pillars: risk mitigation (e.g. masking, sanitation and hygiene, campus de-densification, etc.); viral detection (e.g. asymptomatic and symptomatic testing, environmental monitoring, molecular sequencing); and public health intervention (e.g. isolation/quarantine, contact tracing, digital exposure notification). Environmental monitoring consisted of wastewater monitoring conducted in tandem with diagnostic testing and contact tracing. During the fall term of 2020, approximately 9,700 undergraduate and graduate students lived on the UCSD campus and an estimated 4,000 employees worked on-campus. All on-campus residents were also mandated to participate in the same free and campus-provided diagnostic COVID-19 tests on a biweekly basis, which aided in the validation of the sensitivity and efficacy of the wastewater surveillance. Here, we report findings from our observational study of wastewater monitoring in these high-density buildings, and a cluster randomized study of manholes associated with residential buildings that were randomized to receive wastewater monitors at one of two timesteps. The sensitivity of the monitoring enabled detection of a single asymptomatic individual in a building with 415 residents, thereby illustrating the potential for averting potential outbreaks on campus. Furthermore, quantitative analysis of the wastewater signatures from the isolation units helped glean insight into the micro-scale viral shedding dynamics and its potential implications on the relationship between the viral load in wastewater and positivity rates.

## Implementation of a large-scale wastewater surveillance program

Analyses informed by the Campus GIS were used to identify manholes or sewer cleanouts associated with each residential building on campus. Sixty-eight commercial autosamplers (that can collect time-weighted composite samples) were deployed across the campus residences. Autosamplers were first prioritized to the most proximal manhole to buildings with residential populations greater than 150. This decision was made based on agent-based network modeling of SARS-CoV-2 transmission on the UCSD campus indicating that the highest risk areas for large outbreaks on campus were buildings containing the largest residential populations (10). In parallel to our observational study of wastewater monitoring in these high-density buildings, we additionally performed a cluster randomized study. Clusters of manholes associated with residential buildings were randomized to receive wastewater monitors at one of two timesteps (end of November or end of December). The purpose of this ongoing cluster RCT was to evaluate the impact of wastewater monitoring on outbreak size in the associated buildings. In total across our observational and cluster RCT studies, these 68 autosamplers covered 239 buildings, including the majority of the on-campus residence halls (Fig. 1A). One of the major bottlenecks in the implementation of a large-scale wastewater monitoring system on campus with daily sample collections, is the sample processing time and labor. Here, we employed an automated, high-throughput wastewater processing pipeline which enables the processing of 96 wastewater samples in 4.5 hours (Fig. 1C) reducing the required time, cost and personnel dramatically.

**Figure 1:**
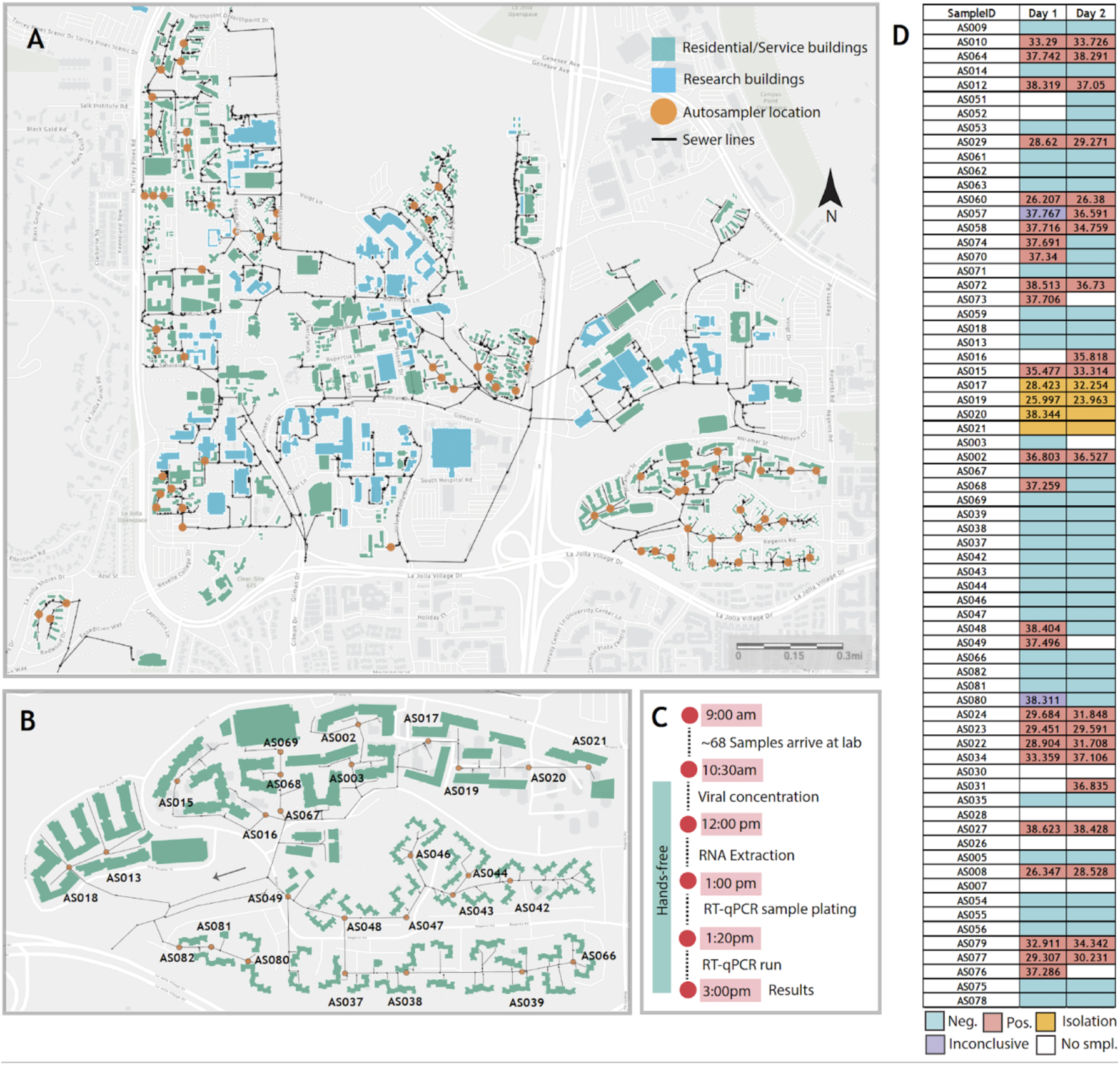
High-throughput wastewater surveillance scheme. A. Map showing the locations of the 68 actively deployed autosamplers (denoted in orange) across the campus residences. B. Snapshot of one of the residence clusters showing the location of 27 samplers covering one of the zones with the highest occupancy on campus. All autosamplers have unique IDs which have been pre-linked to the corresponding manholes on the GIS server. C. Timeline of the daily wastewater sampling and analysis. D. An example of the wastewater sample data reported over two consecutive days. The numbers in the cells indicate the measured cycle threshold values of the N1 gene for the respective sample. Amplification in at least 2/3 genes for both replicates was considered positive. The samplers indicated in yellow were collected from the isolation dorms on campus. Building-specific data have been de-identified in accordance with the University reporting policies.

## Data integration and visualization

Samples from the 68 wastewater samplers were collected daily by field staff. Each autosampler collected wastewater into a sample bottle. Both the autosampler and the sample bottle are associated with a unique scannable barcode which were scanned by the onsite workers using a mobile app when samples were being picked up. The autosamplers were pre-linked to campus asset codes for the manholes they are deployed at for ease of data integration. When pulling samples at particular sites, field staff can easily manage corresponding spatial features using their mobile phones, the samples can be tracked to the lab utilizing the unique ID. The samples were then brought to the laboratory for viral concentration and analyses. SARS-CoV-2 signatures in wastewater was elucidated via RT-qPCR screening for the 3 SARS-CoV-2 specific genes namely, N1, N2 and the E-gene (Fig. 1D). The results for each sample were then pushed into the system, which can be viewed and analyzed by researchers (Fig. 2A). The obtained RT-qPCR results were then integrated with the campus GIS database in order to trace the potential sources of observed positive signals based on which buildings are upstream from an autosampler in the campus sewer network.

**Figure 2:**
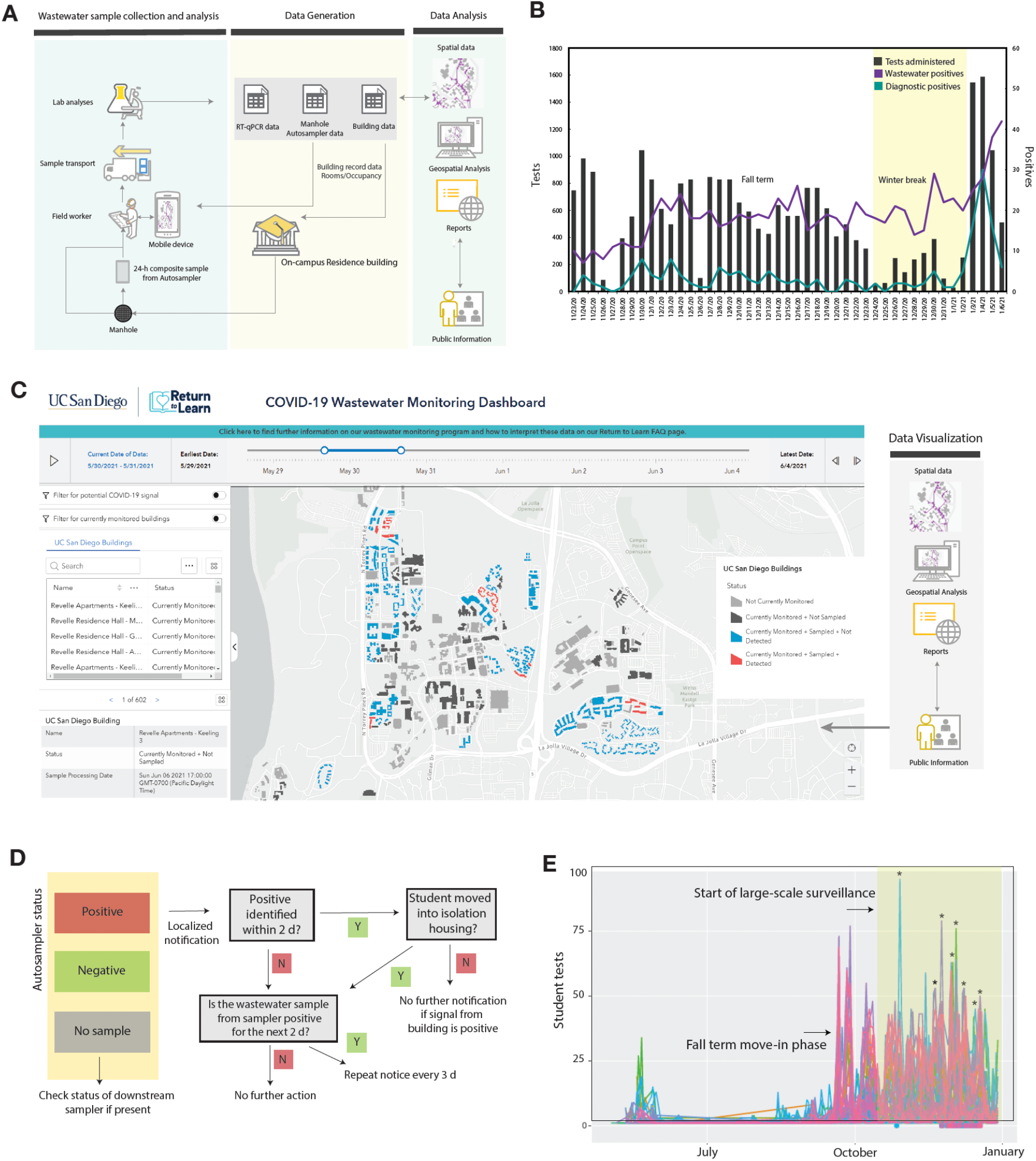
Wastewater surveillance workflow implementation. A. Sample collection to analysis workflow. C. Diagnostic testing and wastewater data shown for a 45-day duration. A spike in the wastewater and subsequently diagnostic was observed prior to the start of the winter term on January 4^th^, 2021 (move-in began on January 1st 2021). C. The interactive public wastewater monitoring dashboard showing the buildings monitored (black) and potential buildings that contributed to a positive wastewater signal (red). The dashboard is updated daily. A slider at the top of the dashboard enables the viewing of historic data. D. Fall quarter 2020 notification process in the case of a positive signal detection from any of the autosamplers. Note: for buildings with public bathrooms, a campus-wide notice was sent. E. Student testing rates associated with each wastewater sampler positive during the study period. The colors represent the individual manholes where the samplers were deployed at and recorded at least one positive during the study period. The dots shown below the x axis indicate that a notification was sent to these students on the corresponding date. Selected testing peaks following wastewater notifications are indicated by asterisks on the plot (further details are provided in Suppl. Table 2).

For instance, if sampler B was positive but an upstream sampler A was negative, only the buildings contributing waste into the sewer between samplers A and B were assumed to be potentially associated with the positive wastewater signal. The spatially-enabled sewer network and subsequent trace of samplers to buildings was stored in and performed by ArcGIS Pro 2.7 (Esri). This data was then visualized by leveraging the ArcGIS Online platform (Esri) into several maps and dashboards. A public-facing interactive wastewater map that displays buildings associated with positive samplers can be viewed at the bottom of the RTL dashboard: https://returntolearn.ucsd.edu/dashboard/index.html (Fig.2C). Other visualizations include internal web maps that display the connection of sewer manholes, cleanouts, and buildings. This workflow increases efficiency and awareness concerning COVID-19 testing and tracing by integrating rich interconnected geospatial and lab results to accurately inform decision making. Targeted email notices were sent to those residing or working in the specific building(s) which were deemed potential sources of the wastewater positive. Campus wide notices were sent (Fig. 2D) in the event a potentially positive building contained a common access area open to the public. The emails alerted individuals to the positive wastewater signal and encouraged individuals to obtain SARS-CoV-2 testing at no charge at any of the testing locations on campus or via the use of self-administered tests available at vending machines across the campus. Additionally, signs were placed in the interior hallways of affected buildings with information on the wastewater positive status.

Wastewater results were corroborated with the diagnostic test results among students living in campus buildings monitored by the wastewater program. During fall 2020, students residing on or coming to campus were mandated to test every other week.

## Data Analytics

### COVID-19 cases identified in relation to wastewater detection and notification

From November 23rd to December 31st, a total of 1574 wastewater samples were collected. Of the collected samples, a total of 692 were positive, 878 negative, and 34 inconclusive. 96 of the total positives obtained were associated with isolation dorms, which served as positive controls. The proportion of samplers collected with a positive signal increased over time (Fig. 2B), consistent with increases in case numbers among residential students and the broader San Diego County over the same time period. Across this period, there were 59 cases diagnosed among on campus students residing in buildings monitored by the wastewater program. Of these, 84.5% (n=50) of these individual case diagnoses were preceded by positive wastewater samples (either in the days prior or the day of diagnostic testing), indicating that the wastewater program was highly sensitive in detecting cases. In only 8% of individual cases (n=5) was the wastewater signal negative the days preceding individual diagnostic detection. 7% (n=4) of individual cases were missed because no sample was obtained the day of or prior to diagnostic detection (Suppl. Table 1). Over the course of our surveillance, 23 cases were identified within 2 days after sending out a localized notice indicating the high overall response rate (Fig. 2E, Suppl. Table 1). The identified students were moved to the designated isolation and quarantine buildings on campus, with the exception of select graduate students who can isolate in place (if living with family). 100 on-campus cases were reported at UC San Diego in the duration of our wastewater surveillance (November 23rd to Dec 31st, 2020). However, 85 new cases were recorded from Jan 1st to Jan 7th, associated with students returning to their on-campus residence after winter break (compared to 11 unique cases associated with students who remained on-campus) (Fig. 2B). During the same period, 435 cases were reported by students who were living off-campus in the San Diego area.

### Impact of wastewater-triggered notifications on testing uptake rates

In order to study the impact of wastewater related notifications and their impact on the testing rates, we analyzed the wastewater data from building(s) associated with individual manholes with notification and test data from those buildings. Data from 36 individual manholes which were associated with wastewater positives (hence targeted notifications) during the period of study were examined. The study period also corresponded to the beginning of the large winter surge in COVID diagnoses in San Diego County, and also with Thanksgiving and Christmas holidays. For each manhole, using the first notification date as the index date, sum of the associated student test numbers over the three days prior to the first notification date was taken as the pre-notification total test number, and the sum of the associated student test numbers on the first notification date plus the two subsequent days as the post-notification total test number. From these pre-post total test numbers formed, two summary statistics were estimated: the difference in test numbers, post-notification minus pre-notification; and the ratio of test numbers, post-notification divided by pre-notification. Across the 36 manholes studied, the median ratio of post-notification test numbers to pre-notification test numbers for the 36 manholes was 1.90 (bootstrap 95% CI 1.48-3.17). The ratio for all but 3 manholes was over 1. The mean difference was 26.7 (normal theory 95% CI 14.6 to 38.7). In almost all cases the notification increased the testing rates 1.5x-13x (Suppl. Table 2, Fig. 2E). Because these data were obtained during a COVID surge, the positive post-pre difference in testing rates might reflect an increasing trend in COVID testing rates during the surge period. Therefore, we also conducted a permutation test to assess whether there was a statistically significant association between a notification event and increased COVID testing rates, controlling for any temporal background changes in testing intensity. With the randomly permuted dates, there is no association between notification and testing, by construction. On the other hand, any general increasing trend (due to notification) during these dates will still be captured in the permuted data. Permutation test p-value is less than 0.001, we reject the null of no association between notification event and increasing in testing.

### Micro-scale viral shedding dynamics

While SARS-CoV-2 signatures in wastewater can be valuable qualitatively, the granularity of its quantitative interpretation still remains to be elucidated, for example to determine the number of individuals in a building and their correlation to the SARS-CoV-2 signal in wastewater. Currently, the viral shedding dynamics and its effects on wastewater are still not well understood on a micro-scale which precludes the quantitative interpretation of such data (11). This is in part, due to the observed wastewater measurement’s dependence on individualized shedding patterns, where the shedding signature itself may be an indication of the onset or severity of infection. The SARS-CoV-2 signatures from isolation dorms were also being monitored on a daily basis in order to study individual viral shedding dynamics over time on a scale ranging from 1-100 infected persons per building. As a simple first step towards understanding these complex shedding dynamics, the aggregate of the collected wastewater signal from all campus samplers was used to find any correlations with the reported on-campus positives. The positive caseload was modeled as the output to an infinite impulse response (IIR) filter with wastewater signal as an input. The resulting IIR filter (essentially the inverse of a filter capturing the shedding dynamics) was used as it was found to be more meaningful to estimate the caseload. Furthermore, the campus sampler network lies on a gravity sewer, where a specific manhole sampling location will be affected by upstream nodes. Although the true shedding signal is expected to be diluted as it flows downstream, we found that a sampler (AS017) placed 276 ft downstream from the sampler at isolation building (AS019), exhibited a positive signal often correlating with the isolation building occupancy (Fig.3A). Further study is required to quantify the persistence of a signal further downstream for any given sampling point. However, the persistence of a signal downstream combined with high probability of a signal erasure, due to the time and flow-weighted discrete wastewater measurements, at any given sampling point could indicate that the true shedding signal at a building is likely represented by a mixture of the collected wastewater signal from multiple associated samplers. Using this data, the transfer function of the filter was estimated using time-discrete data from 11/23 to 1/1 consisting of aggregate daily wastewater data from all campus samplers as the input and reported positive new cases as the output. Figure 3C shows the measured caseload data compared to the predicted filter output, with a 1-day sampling delay (since the samples are 24-h composites). Furthermore, the estimated filter was then applied to the data from the isolation building denoted by AS019, where the wastewater signal was consistently strong due to students in isolation. Over the course of the study, the number of students isolating in these dorms varied between 3-42 (Fig. 3B). Figure 3D shows the measured active caseload compared to the predicted filter output with a 1-day sampling delay. The estimated filter fit the isolation unit data with a correlation coefficient of r = 0.80. This preliminary data highlights the importance of continuing to survey the isolation units in addition to campus wide reporting for learning about the shedding dynamics on campus, and shows that these may be scalable across campus.

**Figure 3:**
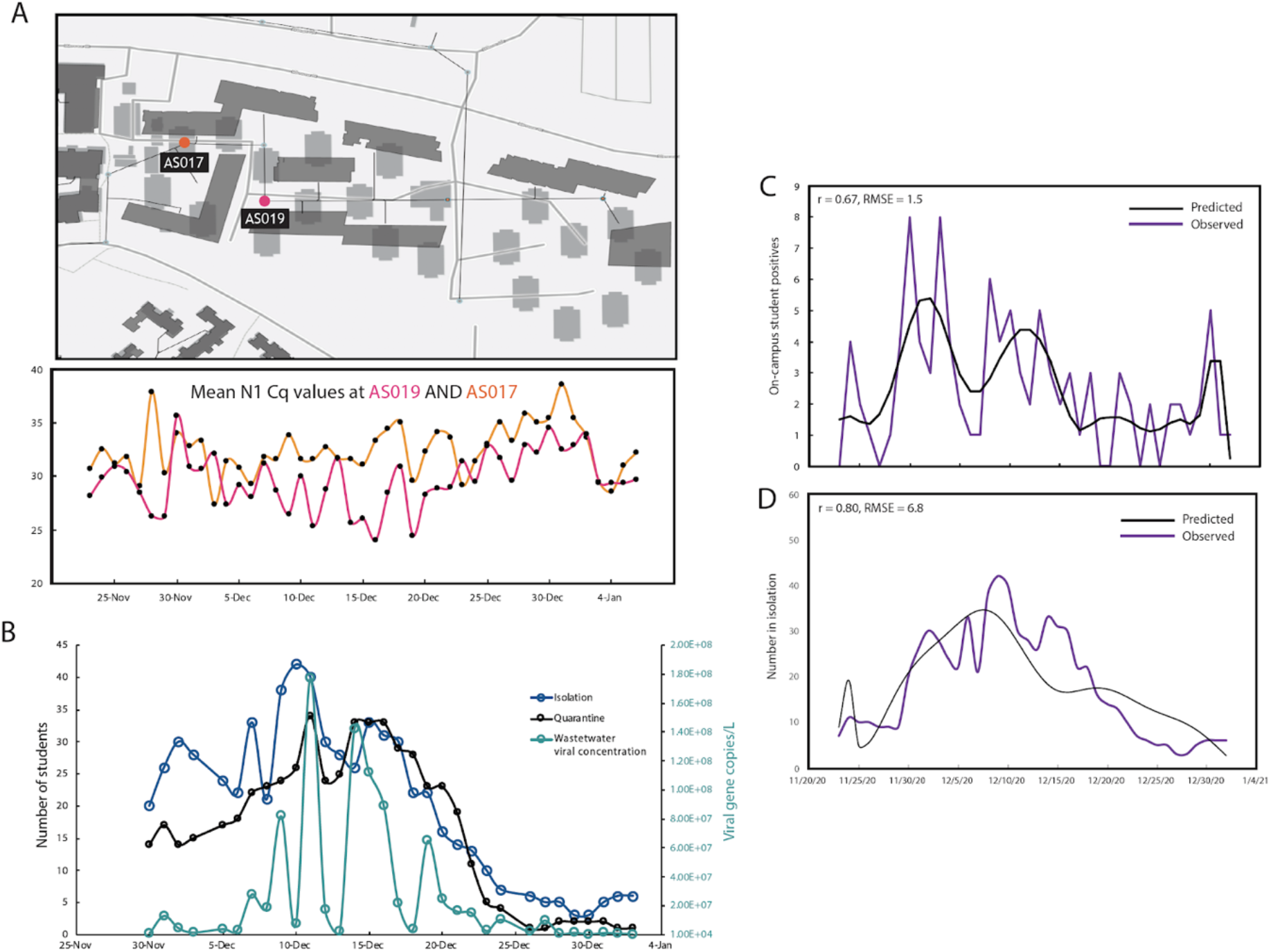
Quantitative interpretation of the wastewater data. A. Snapshot of the sewer network showing the 2 autosamplers by the isolation unit AS017 is downstream of sampler AS019 (associated with the isolation dorm). The mean Cq values of the daily samples from the 2 samplers are shown in the lower panel. B. Mean viral gene copies per L of sewage collected daily from the isolation dorms (sampler AS019) compared to the number students in isolation/quarantine on the same day. C. Measured daily caseload data compared to the predicted filter output with a 1-day sampling delay for all active on campus samplers (mean 0.67, RMSE 1.5). D. Measured daily caseload data compared to the predicted filter output with a 1-day sampling delay for the isolation unit sampler AS019 (mean 0.80, RMSE 6.8).

## Conclusions and future perspectives

Combining the wastewater and observational data can greatly aid in designing optimal intervention strategies using advanced statistical and epidemiological techniques including time series analysis and agent-based modeling. Taken together, our data shows wastewater-based epidemiology can be implemented successfully at the building level, and serve as an important tool for early detection of COVID-19 outbreaks as well as being a cost-effective alternative to high-frequency diagnostic testing. Additionally, it could serve as a longer-term monitoring system when caseloads are lower and only testing a section of the population after encountering a wastewater positive (i.e. responsive testing instead of regular testing). The persistence of a signal in wastewater after a person is no longer infectious (due to extended viral shedding periods in stool) is currently one of the major challenges in wastewater surveillance. This can potentially obscure identification of more asymptomatic cases in the same building. SARS-CoV-2 viral genome sequencing of wastewater can also help resolve these by aiding in elucidating geospatial SARS-CoV-2 genotype distribution. This could also help in early identification of outbreak clusters as well as in tracking newly emerging variants. Currently, the wastewater surveillance program has expanded to cover over 340 campus buildings including the majority of nonresidential buildings and over 15,000 campus wastewater samples have been processed till date.

## METHODS

### Site identification and autosampler deployment

Sampling sites were identified via Campus GIS, which provides mapping and flow direction of interconnected sewer lines and locations of manholes where samplers could be placed. Preliminary dynamic modeling indicated that the largest potential outbreaks would potentially occur within the largest residential buildings, so manholes associated with larger residential buildings were prioritized first.

Large-scale wastewater surveillance across campus began in November with an initial 47 deployed samplers. This number was then increased to 68 by the end of 2020. Autosamplers (HACH AS950) were deployed across the identified sites. All the autosamplers were deployed at manholes and aboveground except 2 which were deployed at sewer ejector pumps due to the inaccessibility of the associated manholes. The manhole covers were modified to enable the passage of the suction tube thereby circumventing the need to install the sampler belowground and the need to open the manhole covers at every sample collection. All the autosamplers were retrofit with 1L Nalgene bottles (Thermo Fisher, Cat# 2104-0032) in order to aid in easy and rapid sample retrieval. All autosamplers were programmed to retrieve samples at 1h intervals over a 24-h period with a pre-and post-purge cycle. The tubing was disinfected after sample retrieval. The pre-barcoded sample bottles were swapped daily. The autosampler barcode and the sample bottle barcodes were scanned by the field staff using the ArcGIS Survey123 mobile app (ESRI) which enabled automatic data integration into the ArcGIS Online environment for trace analysis.

### Sample analysis

The SARS-CoV-2 viral RNA was concentrated from 10ml of raw, unfiltered sewage samples as described elsewhere (1). Briefly, 10 ml of raw sewage was concentrated using an automated affinity-capture magnetic hydrogel particle (Nanotrap) based concentration method using a KingFisher Flex liquid-handling robot platform (Thermo Fisher Scientific, USA). The concentrated viral RNA was then extracted using the MagMAX Microbiome Ultra Nucleic Acid Isolation Kit (Applied Biosystems, Cat # A24357, A24358) using 96 well plates. The RNA was eluted in 50 µL nuclease free water and used for SARS-CoV-2 real time RT-qPCR. The RT-qPCR reactions were carried out in a CFX384 Real-Time System (BIO-RAD) thermocycler in 384 well plates for 3 gene targets (N1/N2/E-gene). qPCR Sample plating was performed using an EpMotion automated liquid handler (Eppendorf, Germany). 5-fold serial dilutions of the positive controls in nuclease-free water and wastewater sample extracts were run to assess inhibition. In addition, an internal amplification control (IAC) was included in every run to control for PCR inhibition. Positive controls and extraction blanks were included in every run. A synthetic RNA encoding the E and N genes of SARS-CoV-2 was used as the positive control (Promega Corp., Cat.no. CS317402). A ladder of 6-fold dilutions were run with every qPCR run.. The no-template control (NTC) was nuclease-free water Amplification of 2/3 genes was regarded as positive, while 1/3 was regarded as inconclusive (only Cq values <40 in all targets were considered positive). Pepper mild mottle virus (PMMoV) was also screened to adjust for daily load changes. The Cq values for the N1 gene ranged from 21.121 to 38.513 for the wastewater samples with an average of 33.198 for the samples measured in the duration of study. The average Cq values for the isolation dorms were 29.067 (SD 2.52). Recovery experiments were conducted by spiking in serial dilutions of heat-inactivated SARS-CoV-2 viral particles into raw sewage samples. Detailed sampling to analysis protocol is available at: dx.doi.org/10.17504/protocols.io.bshvnb66

### Automated Data reporting

The wastewater data reporting is automated by AUM (*auto-update microservice*), a microservice hosted by AWS Lambda, through a docker image for all the dependencies, such as Google Sheet API. Each day when the raw Cq values get uploaded, the microservice automatically converts the Cq values to a format that can be cross referenced with the plate map and automatically update the data report on a Google Sheet that has the longitudinal wastewater data across all deployed samplers. The data from the sheet is then linked to the GIS server to automatically update the dashboard thereby streamlining the data integration process. The detailed architecture is available at the Github repository for replication and deployment. https://github.com/CrisZong/AUM.git

## Data Availability

Relevant links to data accessions have been provided in the main manuscript.

## ACKNOWLEDGEMENTS

The authors would like to thank UC San Diego’s Return to Learn (RTL) program for funding the campus-wide wastewater surveillance efforts. The authors would also like to thank Robert M. Neuhard and the rest of the RTL leadership team and Bradley Sollenberger of the Operational Strategic Initiatives (OSI) team for ensuring the smooth functioning of the program along with the EXCITE (EXpedited COVID-19 IdenTification Environment) CLIA lab at UCSD and the SEARCH (San Diego Epidemiology and Research for COVID Health) Alliance for processing all campus diagnostic tests. We also thank Jason Kayne, Rich Cota, Jesus Ortiz and the Facilities management team (FM) at UCSD, Joseph Mayer from the Center for Aerosol Impacts on Chemistry of the Environment (CAICE) and Luke Arnold of the Campus Research Machine Shop (CRMS) for assistance with the installation and operation of the autosamplers; Robbie Jacobs, Shawn Knepple and their team at UCSD Logistics for assisting with our daily sampling efforts; Dr. Christopher Longhurst, MD and the UC San Diego Health Information Services team; Brett Pollak and the UCSD Information Technology Services team for assisting with the daily notifications; Alysson M. Satterlund, Angela Song, Angela Scioscia and Elizabeth H. Simmons of Academic Affairs for contact tracing and targeted campus messaging assistance; Jana Severson, Patrick Hochstein, Hemlata Jhaveri and the UCSD HDH team and the UCSD Environmental Health and Safety (EHS) personnel; Dr. Jack Gilbert and the Microbiome Sample Processing Core at UC San Diego for access to qPCR equipment.

