## Supplemental Data for "Rapid, large-scale wastewater surveillance and automated reporting system enabled early detection of nearly 85% of COVID-19 cases on a University campus"

**Supplemental Table S1:** **Categorization of COVID-19 cases in relation to wastewater sampling, detection, and notification occurring between November 23, 2020 and December 31, 2020.**

| Diagnoses of COVID-19 cases in relation to wastewater detection and notification | | **n** | **%** |
| --- | --- | --- | --- |
| Positive signal preceding diagnosis | Case diagnosed **less than 3 days** from a notification | 23 | 39% |
|  | Case diagnosed **more than 3 days** from a notification | 7 | 12% |
|  | Case diagnosis coincident with positive signal (no notification) | 5 | 8% |
|  | Detected but downstream from isolation building (no notification) | 4 | 6% |
|  | Detected but notification occurred the day after case diagnosed because of notification delay | 5 | 8% |
|  | Case diagnosed where there was a known isolation (no notification) | 6 | 10% |
|  | Case diagnosed but not detected by wastewater | 5 | 8% |
|  | Sample not collected and no notification | 4 | 7% |
|  | **Total cases diagnosed** | **59** | **100%** |

**Supplemental Table S2: Testing uptake rates in buildings associated with each manhole pre- and post-notification after a wastewater positive**

| **Manhole ID** | **Notification Date** | **Post-notification tests** | **Pre-notification tests** | **Difference** | **Ratio** |
| --- | --- | --- | --- | --- | --- |
| C1M031 | 10/27/20 | 100 | 49 | 51 | 2.0408 |
| C1M057 | 11/25/20 | 33 | 46 | -13 | 0.7174 |
| C1M059 | 12/6/20 | 86 | 88 | -2 | 0.9773 |
| C1M060 | 12/4/20 | 31 | 39 | -8 | 0.7949 |
| C1M161 | 12/6/20 | 67 | 66 | 1 | 1.0152 |
| C2M015 | 11/24/20 | 51 | 33 | 18 | 1.5455 |
| C3M022 | 12/13/20 | 7 | 3 | 4 | 2.3333 |
| C3M149 | 11/29/20 | 20 | 5 | 15 | 4 |
| C3M158 | 12/18/20 | 52 | 23 | 29 | 2.2609 |
| C3M159 | 11/29/20 | 27 | 3 | 24 | 9 |
| C6M021 | 12/5/20 | 29 | 25 | 4 | 1.16 |
| C6M025 | 12/1/20 | 157 | 58 | 99 | 2.7069 |
| C6M030 | 12/8/20 | 6 | 1 | 5 | 6 |
| C6M033 | 12/3/20 | 35 | 11 | 24 | 3.1818 |
| C6M034 | 12/3/20 | 25 | 3 | 22 | 8.3333 |
| C6M041 | 12/8/20 | 14 | 10 | 4 | 1.4 |
| C6M042 | 12/8/20 | 6 | 4 | 2 | 1.5 |
| C6M043 | 12/13/20 | 10 | 2 | 8 | 5 |
| C6M045 | 12/4/20 | 16 | 8 | 8 | 2 |
| C6M046 | 11/24/20 | 24 | 7 | 17 | 3.4286 |
| C6M047 | 11/24/20 | 5 | 3 | 2 | 1.6667 |
| C6M049 | 12/8/20 | 10 | 8 | 2 | 1.25 |
| C6M072 | 10/27/20 | 145 | 39 | 106 | 3.7179 |
| C6M092 | 11/24/20 | 61 | 34 | 27 | 1.7941 |
| C7M005 | 11/28/20 | 130 | 41 | 89 | 3.1707 |
| C7M008 | 11/28/20 | 12 | 1 | 11 | 12 |
| C7M012 | 11/24/20 | 121 | 87 | 34 | 1.3908 |
| C7M017 | 11/29/20 | 146 | 11 | 135 | 13.2727 |
| C7M025 | 12/9/20 | 18 | 14 | 4 | 1.2857 |
| C7M027 | 11/24/20 | 80 | 59 | 21 | 1.3559 |
| C7M030 | 12/3/20 | 35 | 24 | 11 | 1.4583 |
| C7M031 | 12/1/20 | 56 | 37 | 19 | 1.5135 |
| C7M032 | 12/1/20 | 17 | 11 | 6 | 1.5455 |
| S2M016 | 12/18/20 | 7 | 5 | 2 | 1.4 |
| SMISC-079 | 11/29/20 | 127 | 21 | 106 | 6.0476 |
| SMISC-080 | 11/29/20 | 91 | 18 | 73 | 5.0556 |
